# The textures of sarcoidosis: quantifying lung disease through variograms

**DOI:** 10.1101/2024.05.20.24307618

**Authors:** William L. Lippitt, Lisa A. Maier, Tasha E. Fingerlin, David A. Lynch, Ruchi Yadav, Jared Rieck, Andrew C. Hill, Shu-Yi Liao, Margaret M. Mroz, Briana Q. Barkes, Kum Ju Chae, Hye Jeon Hwang, Nichole E. Carlson

## Abstract

**Objective:** Sarcoidosis is a granulomatous disease affecting the lungs in over 90% of patients. Qualitative assessment of chest CT by radiologists is standard clinical practice and reliable quantification of disease from CT would support ongoing efforts to identify sarcoidosis phenotypes. Standard imaging feature engineering techniques such as radiomics suffer from extreme sensitivity to image acquisition and processing, potentially impeding generalizability of research to clinical populations. In this work, we instead investigate approaches to engineering variogram-based features with the intent to identify a robust, generalizable pipeline for image quantification in the study of sarcoidosis.

**Approach:** For a cohort of more than 300 individuals with sarcoidosis, we investigated 24 feature engineering pipelines differing by decisions for image registration to a template lung, empirical and model variogram estimation methods, and feature harmonization for CT scanner model, and subsequently 48 sets of phenotypes produced through unsupervised clustering. We then assessed sensitivity of engineered features, phenotypes produced through unsupervised clustering, and sarcoidosis disease signal strength to pipeline.

**Main results:** We found that variogram features had low to mild association with scanner model and associations were reduced by image registration. For each feature type, features were also typically robust to all pipeline decisions except image registration. Strength of disease signal as measured by association with pulmonary function testing and some radiologist visual assessments was strong (optimistic AUC ≈ 0.9, *p* ≪ 0.0001 in models for architectural distortion, conglomerate mass, fibrotic abnormality, and traction bronchiectasis) and fairly consistent across engineering approaches regardless of registration and harmonization for CT scanner.

**Significance:** Variogram-based features appear to be a suitable approach to image quantification in support of generalizable research in pulmonary sarcoidosis.

## 1. Introduction

Sarcoidosis is a systemic granulomatous disease affecting the lungs in over 90% of patients (Baughman et al. 2011). The disease is highly heterogeneous in presentation and prognosis, and etiology and treatment of disease remain under investigation (Baughman & Lower 2015). Sarcoidosis often presents in the lungs as textural changes in the lung parenchyma. In practice, these abnormalities may be qualitatively assessed on chest computed tomography (CT) scans by radiologists to produce visual assessment scores (VAS) for presence and severity of abnormalities with different visual textures. Qualitative visual assessment has been shown to have low inter- and intra-rater reliability (Van den Heuvel et al. 2015, Lovinfosse et al. 2022, Benn et al. 2024). Reliable quantification of disease related textures from images could support ongoing efforts to identify meaningful phenotypes of pulmonary sarcoidosis (Desai et al. 2023, Carlson et al. 2024, Lin et al. 2022, Schupp et al. 2018, Lew et al. 2023, Rubio-Rivas & Corbella 2020).

A standard approach to quantify texture or spatial structure in images is radiomics (Van Timmeren et al. 2020, Haralick et al. 1973). In radiomics-based approaches, statistical measures of texture are computed from images and these measures serve as observations of engineered features in later analyses. Previous investigation of whole-lung classical radiomics in sarcoidosis showed consistently strong associations with clinical measures of disease such as pulmonary function testing (PFT) (Carlson et al. 2024, Ryan et al. 2019). However, computed radiomic feature values are known to vary substantially on the bases of differences in image acquisition, reconstruction, and processing (Rizzo et al. 2018, Mackin et al. 2015, Van Timmeren et al. 2020, Shiri et al. 2020). Indeed, in radiomics applications, the scanners which were used to obtain images may account for more variability than disease, necessitating harmonization (Rizzo et al. 2018, Mackin et al. 2015). Although harmonization or batch correction can be useful to produce meaningful analyses within a single study, harmonization approaches are typically not generalizable beyond the particular application, leaving it difficult to move these measures into less controlled settings such as clinical use. This suggests we might improve upon previous work by identifying alternative approaches to feature engineering which efficiently capture sarcoidosis-relevant textures while disregarding systematic variation due to study factors unrelated to disease.

In this work, we investigate potential clinical relevance of and optimal pipelines for engineering variogram-based features in the study of pulmonary sarcoidosis, with specific interest in exploring phenotyping via unsupervised clustering. Variograms are a common geostatistical tool for measuring and modelling spatial covariance within an image or spatial process as a function of distance between locations (Banerjee et al. 2014). Empirical variograms have previously been used as a type of engineered radiomic feature in biomedical imaging to investigate health associations with spatial structures like texture, such as associating age and sex with bony trabecular shadows (Gough et al. 1994), investigating chromatin patterns in various tissues (Diaz et al. 1997, Muniandy & Stanslas 2008), assessing malignancy of lung nodules (Silva et al. 2004), and identifying the presence and location of tumors in the breasts (Ericeira et al. 2013). We did not find previous application of variograms in the study of interstitial lung diseases like sarcoidosis, though other measures specifically related to spatial covariance such as Geary’s C and Moran’s I have been considered (Ryan et al. 2019). Models for variogram functions typically include overall magnitude of variation and a notion of spatial scale of correlation as parameters, and tend to be more interpretable measures of spatial structure than their classical radiomic counterparts. As variograms are closely related to measures of spatial covariance, variogram features also have the potential to be less sensitive to noise and distortions attributable to image acquisition which happen not to affect the spatial covariance structure in particular.

Use of variogram-based features as engineered radiomic features is historically handled differently from classical radiomics, for which robustness of features to the imaging pipeline is considered fundamentally important (Van Timmeren et al. 2020, Zwanenburg et al. 2020, Lambin et al. 2017). While empirical variogram features have long been observed to also differ with image acquisition and pre-processing factors, these features have been valued as a sensitive analytic tool for that reason (Carr & De Miranda 1998, Goodin et al. 2004, Dai & Khorram 1998, Luo et al. 2022). Investigations of variograms as radiomics routinely consider optimal decisions such as regions and resolutions for computation and standardization of images with respect to image contrast (Gough et al. 1994, Diaz et al. 1997, Keil et al. 2012, Jacob et al. 2013, Jacob & Carson 2014). By comparison, relatively little attention has been given to classical radiomics as a sensitive tool for which image processing and feature engineering might be optimized to produce strong signal (Au et al. 2021, Lv et al. 2018). While robustness of feature computation to variability in image acquisition and processing is desirable in that it supports generalizability of research to new cohorts and contexts, understanding which processing decisions lead to efficient extraction of disease signal is also fundamentally important.

The purpose of this paper is to establish clinical relevance of and identify an appropriate pipeline for variogram-based study of sarcoidosis chest CT phenotypes in future clinical cohorts. To uncover influential decisions in the engineering pipeline, we quantified sensitivity of variogram-based features and clusters to processing specifications under 24 different feature engineering pipelines producing 48 clustering analyses. We further assessed presence and strength of disease-relevant signals of heterogeneity for each clustering analysis to identify pipelines which best supported study of sarcoidosis.

## 2. Materials and Methods

### 2.1. Data

The Genomic Research in Alpha-1 Antitrypsin Deficiency (AATD) and Sarcoidosis (GRADS) study (Moller et al. 2015, Vukmirovic et al. 2021) was a multi-site study in the United States (N=368). All subjects gave written informed consent according to the site’s institutional review board. Enrolled subjects underwent physical examination, pulmonary function testing (PFT), research chest X-ray (CXR), and high resolution computed tomography (CT) imaging.

CXR was performed based on the site’s standard protocol and Scadding Stage was determined by the site radiologist. CT was obtained in accordance with the GRADS protocol (Moller et al. 2015). 3D chest CT scans were obtained as a series of 2D axial images or slices. Scans from this cohort included a complete set of sequentially adjacent axial slices.

From the GRADS cohort, N = 337 subjects had CT scans available and of sufficient quality for investigation in this study; that is, inspiratory scans of appropriate resolution and reconstruction lacking artifacts and successfully masked and segmented. See (Carlson et al. 2024) for details of original image acquisition, reconstruction, and selection for use in analysis. All images were resampled to have the same resolution (1 x 1 x 1 mm) and centered. Images were then masked and segmented using the `lungct` package (Ryan et al. 2020) in R (R Core Team 2020), producing two binary images indicating presence of left or right lung tissue. For registered analyses, an individual’s 3D lung mask was further registered to a sarcoidosis-specific template lung mask, and the corresponding transformation was then used to register each individual’s CT scan to a template lung. See (Ryan et al. 2020) for details of masking and registration.

While GRADS CTs were interpreted for many CT abnormalities by a dedicated thoracic chest radiologist determined by GRADS to evaluate sarcoidosis and AATD, we used a simplified visual assessment scoring (VAS) method developed by our radiologists for sarcoidosis. Experienced thoracic radiologists evaluated CT for the presence of the following features: mediastinal and hilar lymphadenopathy, micronodules, ground-glass opacity, consolidation, conglomerate perihilar or peribronchovascular mass, linear or reticular abnormality, fibrotic abnormality, honeycombing, mosaic attenuation, architectural distortion, traction bronchiectasis, bronchial wall thickening, cysts, cavity, emphysema, and air trapping. Definitions of these findings were based on (Bankier et al. 2024). For this work, we considered only a subsetted VAS panel for abnormalities deemed present in 15-85% of the GRADS cohort: mediastinal and hilar lymphadenopathy, micronodules, ground-glass, conglomerate mass, reticular abnormality, fibrotic abnormality, mosaic attenuation, architectural distortion, and traction bronchiectasis.

PFT was performed according to established criteria per the GRADS protocol (Moller et al. 2015), including post broncho-dilation forced expiratory volume in one second (FEV1), forced vital capacity (FVC), ratio of FEV1 to FVC (FEV1/FVC), and single-breath carbon monoxide diffusing capacity (DLCO). DLCO was corrected for elevation in accordance with (Graham et al. 2017).

Collected demographics included age, height, weight, BMI, sex, and self-reported primary race and ethnicity. Self-reported primary race and ethnicity were collapsed in this study into a single marker to reduce identifiability of participants with rarer self-reported descriptions and improve power in analyses as follows: all subjects reporting Hispanic ethnicity were reported in this study as Hispanic subjects and all subjects self-reporting ethnicity as non-Hispanic and primary race as White or as Black were reported as White subjects or as Black subjects respectively. All remaining subjects self-reported a primary race as Asian, American Indian, or Alaska Native, or did not identify a single primary race (identifying as multi-racial, having no primary race, or having unknown primary race), and are reported together as subjects in a combined group.

#### 2.1.1. Cohort descriptive statistical analysis

Covariates comprised of demographics age, height, sex, BMI, and race/ethnicity, PFT outcomes, and presence of VAS outcomes are summarized for the N = 337 individuals in this study, stratified by Scadding stage, and reported with counts and percents for categorical covariates and with means and standard deviations for continuous covariates.

### 2.2. Feature engineering

#### 2.2.1. Variogram Theory

We discuss variogram theory explicitly in the context of 3D images such as CT scans. In this context, a voxel is a 3D analog of the 2D pixel. To describe the models, we will assume stationarity, meaning the image has the same distribution at every location, and isotropy, meaning the image has the same distribution in every direction. As described below, we can later relax these assumptions for investigation of their impact on estimation.

Under further reasonable assumptions, the variogram function *γ*(*h*) may be understood as *C*(0) *− C*(*h*) where C(*h*) is the common covariance of the hues of any two voxels separated by a distance *h*. More generally, let *z* denote an image meeting the above conditions with hues *z* = {*z_i_*}_*i*∈*I*_ at observed voxels I. If *z* is observed at any two voxels *i, j* ∈ *I* a distance *h* apart, we may define

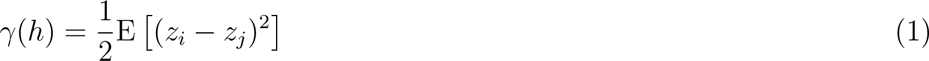

noting this definition to be independent of the specific pair (*i, j*) by stationarity and isotropy. See (Banerjee et al. 2014) for more detail. We note that terminology around the variogram is not settled. Some might say semivariogram or semivariance (Banerjee et al. 2014) or *γ*-variance (Bachmaier & Backes 2011), among other terms. We use the term variogram for limited consistency with established literature.

To model variograms, we consider the Matérn family as well as the special case of the exponential model. The Matérn family of models is very flexible, standardly used, and mathematically natural for variograms (Banerjee et al. 2014). The Matérn family has parameters nugget, partial sill, range, and kappa or degrees of freedom. In turn, these parameters may be understood to describe noise, overall variability not attributed to noise, the distance at which image voxels appear to become roughly uncorrelated, and image smoothness. Exponential models are a widely used special case which are more analytically tractable with kappa (image smoothness) set to 0.5.

For a high resolution image of a large region such as chest CT of the lungs, the variogram value *γ*(*h*) is well-estimated empirically using voxel pairs a distance *h ± ɛ* apart. If we expect the assumption of stationarity to be violated by a non-constant mean, empirical variograms may also be computed from residuals obtained by subtracting off an assumed or estimated mean image from the observed image. In absence of statistical models, variogram models are classically fit from empirical variograms using weighted least squares (Cressie 1985).

#### 2.2.2. Application to GRADS

From the chest CT scans, 24 similar datasets were engineered through the use of variograms; see Figure 1. Datasets were distinguished through the decisions to a) register images to a template or not, b) assume stationarity or account for linear drift in computation of empirical variograms, c) use empirical variograms or fit variogram model parameters for either exponential or Matérn models, and d) harmonize or not harmonize data for CT scanner effects; details follow.

**Figure 1.**
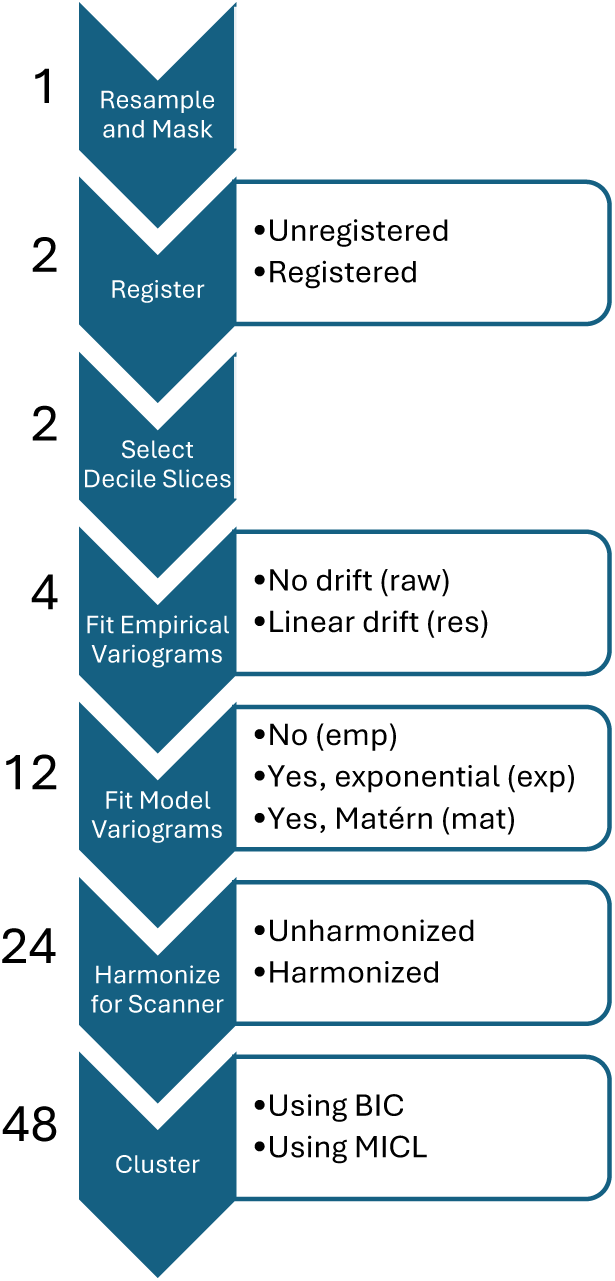
Steps in the image processing, feature engineering, and clustering pipeline. The number of datasets or analyses after completing each step is listed to the left, and steps permitting decisions have decision options listed to the right.

To ensure sufficient data for variogram estimation, all top and bottom axial slices of the left lung and right lung images with fewer than 1000 voxels indicated to be lung tissue by the corresponding mask were removed from consideration. Unregistered images paired with the unregistered mask or registered images paired with the template mask were then used for this study. For unregistered images, this typically produced images with 150-300 axial slices with at most 3.3% of lung voxels removed from consideration. For registered images, this produced 228 (left) and 232 (right) sequential axial slices with 0.3% of lung voxels removed from consideration.

The number of axial slices considered was then substantially reduced to account for the sometimes reduced availability of axial slices in clinically obtained images. Requiring fewer slices supports broader inclusion of participants and potentially more consistent feature extraction. Specifically, for each image, observations for the inner decile axial slices of each lung were extracted, resulting in 18 axial slices per 3D CT image. That is, for each lung, 9 axial slices were selected which broke the masked lung into 10 equal height segments. Decile 1 corresponded with the lowest retained axial slice of the lung while decile 9 corresponded with the uppermost retained axial slice. Decile indices were determined independently for individuals and for right and left lungs. These decile slices were then used for computation of variogram features.

For each axial slice, two empirical variograms were computed under the assumption of either stationarity and isotropy (from raw data) or the same except permitting a linear mean trend (from linear residuals). The `variogram` function from the `gstat` package (Gräler et al. 2016) in R was used for computation with a cutoff of 25mm, a formula of `HU ∼ 1` indicating no drift, and otherwise default settings for variograms from raw data. Default settings estimated variogram values at 15 *h* values from 0 to the cutoff using equal size bins. For variograms from linear residuals, the same function was used with a cutoff of 25mm, a formula of `HU ∼ x + y` indicating linear drift, and otherwise default settings. This produced four empirical variogram datasets differing by decisions on registration and accounting for drift.

An additional eight engineered datasets were computed from these four by fitting either an exponential or Matérn variogram model to the empirical variograms and using the fit model parameters as features. Models were fit using the `fit.variogram` function from the `gstat` package in R with initial `psill` value given by the maximum empirical variogram value, initial `range` value 1, and `fit.method` 2 corresponding with weighted least squares. In each case, the nugget parameter (noise) is set to zero. For computational reasons, parameter kappa is not fit continuously but rather permitted to take one of a list of pre-specified values: `seq(0.01,5,0.01)`. Fit range and psill parameters were then log-transformed to account for skew and standardized. For two subjects, model fit parameters were estimated as negative for some datasets, which is not reasonable. These two subjects were thus removed across all datasets to preserve comparability, resulting in N = 335 subjects with unharmonized engineered feature data.

Finally, from these twelve datasets, another twelve datasets were produced via harmonization. The `ez.combat` package (Koscik 2021) in R was used with default settings to adjust for CT scanner model while preserving variability associated with Scadding stage, height, age, BMI, and sex. N = 330 subjects had complete Scadding and demographic data for producing harmonized engineered feature data.

In summary, 24 datasets were computed, differing by registration, accounting for drift, data type (empirical, exponential, Matérn), and harmonization; see Figure 1. For each dataset, lung-deciles were presumed to correspond across individuals, resulting in balanced datasets with features identified by lung, decile, and value computed, either an empirical variogram value at a specific distance *h* (*p* = 270 features) or an estimated parameter value for a specific variogram model parameter (*p* = 36 features for exponential model, *p* = 54 features for Matérn model).

#### 2.2.3. Feature robustness and association with scanner

We assessed dataset association with scanner model by regressing each feature in each dataset onto scanner model and reporting dataset distributions of model R squared and F-test p-value.

We also assessed robustness or stability of feature computation for matched pairs of variables across datasets. For a pair of variables from different datasets, the variables were considered matched if they were computed for the same lung-decile and were either (a) both empirical variogram values at the same specified distance or (b) both estimates of the same parameter. For example, the right lung decile 9 empirical variogram values at distance 25mm would be matched for the (empirical, linear residual, unregistered, unharmonized) dataset and the (empirical, raw data, unregistered, harmonized) dataset, as would the left lung decile 1 estimates of partial sill for the (exponential, linear residual, unregistered, harmonized) dataset and the (Matérn, raw data, registered, unharmonized) dataset.

Following (Lv et al. 2018, Denzler et al. 2021), for each matched pair of variables, for all available data (N = 330, 335) we computed the two-way random effects, single measurement intra-class correlation coefficient for consistency (ICC(3,1)) using the `icc` function from the `irr` package (Gamer et al. 2019) in `R` with specifications `model="twoway"`, `type="consistency"`, `unit="single"`. In accordance with (Koo & Li 2016, Lv et al. 2018, Denzler et al. 2021), we understand ICC values above 0.9 to indicate excellent robustness.

We compute also the Spearman’s correlation coefficient using the cor function in R to investigate monotonicity of association between matched pairs in absence of robustness.

### 2.3. Clustering analyses

#### 2.3.1. Clustering

Each of the 24 datasets was clustered in two ways (unharmonized data: N = 335; harmonized data: N = 330), producing 48 clustering analyses; see Figure 1. Model-based clustering was used, specifically a sparse diagonal Gaussian mixture model assuming homoscedasticity of irrelevant features and selecting sparsity using an information criterion (Celeux & Govaert 1995, Marbac & Sedki 2017). The `VarSelCluster` function from the `VarSelLCM` package (Marbac & Sedki 2017, Marbac et al. 2020) in R was used to cluster features allowing for between 1 and 8 resultant clusters. One clustering approach used default settings which performs model selection using the Bayesian Information Criterion (BIC). The other clustering approach used Maximum Integrated Complete-data Likelihood (MICL) for model selection and otherwise default settings. The MICL is a less standard information criterion previously observed to outperform BIC when paired with the VarSelLCM approach (Marbac & Sedki 2017). Features selected as relevant and number of clusters selected in cluster analysis are reported.

As we use empirical and model fit variogram measures which are highly related to measures of covariance as features for clustering, we note that we effectively take a covariance-based clustering approach (Hallac et al. 2017, Ieva et al. 2016) similar to that of (Marquez et al. 2021).

#### 2.3.2. Cluster descriptives and comparisons

Each cluster analysis was compared in turn to covariates scanner model, PFT, and VAS outcomes. Cluster analyses were further compared pairwise to each other to assess sensitivity of clustering results to data processing and analysis decisions. Strength of association was assessed using R squared values when comparing to PFT and bias-corrected Cramér’s V (Bergsma 2013) otherwise. Differences in covariates and in demographics across groupings were assessed and p-values reported via Fisher exact test with simulated p-value from 20,000 replicates for categorical covariates or via F-test p-value for continuous covariates. For these analyses, complete case analyses were performed for each assessment individually.

#### 2.3.3. Cluster associations with clinical outcomes

We assessed strength of association between clusters and clinical outcomes PFT and VAS after adjusting for demographics or for demographics and Scadding stage. Linear regression was used to predict PFT and Firth logistic regression was used to predict VAS. Demographics adjusted for included sex, height, age, and BMI. Note, we exclude self-reported race and ethnicity as an adjustment factor in these analyses. The nature of the relationship between self-reported race and ethnicity and disease presentation in sarcoidosis remains under study (Hena 2020, Zhou et al. 2021, Sharp et al. 2020), and adjustment could hide disease signal or compound systematic biases in selection of appropriate pipelines.

For each outcome, two models were fitted without clusters, one with covariates demographics (base model) and one with covariates demographics and Scadding stage (Scadding only model). Then, for each cluster analysis, two more models were fitted, one with covariates demographics and clusters (cluster only model), and one with covariates demographics, clusters, and Scadding stage (full model).

For each model predicting PFT, we reported an R squared value. For each model predicting VAS, we reported an AUC value obtained from predicting the observed outcomes. Models were fitted to and AUC values were computed for the same data, and so AUC values are ‘optimistic’ training AUC values and understood here as a summary of model fit. For each pair of outcome and cluster analysis, we report likelihood ratio test p-values (PFT) or penalized likelihood ratio test p-values (VAS) for 3 pairs of models: the full and Scadding only models, the cluster only and base models, and the full and cluster only models. Additionally, for each outcome, we report a p-value comparing the Scadding only and base models.

For these analyses, subjects with complete adjustment demographic, Scadding stage, and PFT data (N = 313) were analysed for PFT outcomes and subjects with complete adjustment demographic, Scadding stage, and VAS data (N = 322) were analysed for VAS outcomes.

## 3. Results

### 3.1. Cohort descriptives

Subjects were 54% Female and 53 years old on average; see Table 1 (N = 337). Subjects predominantly identified as Non-Hispanic and White (69%) followed by Non-Hispanic and Black (24%). All PFT and VAS assessments appear to differ substantially by Scadding stage with subjects in stage 0 exhibiting least severe disease and subjects in stage IV exhibiting most severe disease as expected. Inconsistencies between CT-based assessments (VAS) and CXR-based assessments (Scadding) of lymphadenopathy, parenchymal abnormalities, and fibrosis have been previously noted (Zhang et al. 2022), including for this cohort for lymphadenopathy and parenchymal abnormalities (Benn et al. 2024). Inconsistencies included 46% and 32% of subjects in stages II and III having fibrotic abnormalities on CT.

**Table 1.** Demographics, post-bronchodilator PFT, and VAS by Scadding for full cohort.

|  | N<br>miss | Scadding Stage |  |  |  |  |  | Overall, N = 337 |
| --- | --- | --- | --- | --- | --- | --- | --- | --- |
|  |  | 0, N = 43 | I, N = 69 | II, N = 97 | III, N = 45 | IV, N = 79 | NA, N = 4 |  |
| **Sex** | 2 |  |  |  |  |  |  |  |
| Female |  | 27 (62.8%) | 43 (62.3%) | 43 (44.3%) | 23 (51.1%) | 43 (55.1%) | 2 (66.7%) | 181 (54.0%) |
| Male |  | 16 (37.2%) | 26 (37.7%) | 54 (55.7%) | 22 (48.9%) | 35 (44.9%) | 1 (33.3%) | 154 (46.0%) |
| **Race/Ethnicity** | 3 |  |  |  |  |  |  |  |
| White |  | 33 (76.7%) | 54 (78.3%) | 65 (67.0%) | 30 (68.2%) | 44 (56.4%) | 3 (100.0%) | 229 (68.6%) |
| Black |  | 6 (14.0%) | 12 (17.4%) | 22 (22.7%) | 12 (27.3%) | 28 (35.9%) | 0 (0.0%) | 80 (24.0%) |
| Hispanic |  | 3 (7.0%) | 3 (4.3%) | 4 (4.1%) | 1 (2.3%) | 5 (6.4%) | 0 (0.0%) | 16 (4.8%) |
| Combined* |  | 1 (2.3%) | 0 (0.0%) | 6 (6.2%) | 1 (2.3%) | 1 (1.3%) | 0 (0.0%) | 9 (2.7%) |
| **Age (years)** | 2 | 53.04 (9.51) | 51.57 (11.47) | 53.15 (9.29) | 50.62 (10.89) | 55.16 (9.02) | 53.04 (5.86) | 52.94 (9.99) |
| **BMI (kg/m<sup>2</sup>)** | 0 | 33.40 (6.46) | 31.81 (6.93) | 29.53 (5.38) | 32.49 (7.35) | 28.29 (5.57) | 31.45 (6.97) | 30.62 (6.42) |
| **Height (in)** | 0 | 66.67 (4.08) | 66.96 (3.45) | 67.71 (4.23) | 66.49 (4.23) | 66.51 (4.04) | 63.75 (4.19) | 66.93 (4.03) |
| **DLC<sub>0</sub> (ml/min/mmHg)** | 18 | 24.45 (7.52) | 23.86 (7.82) | 23.69 (7.39) | 21.54 (8.06) | 17.60 (7.16) | 20.80 (1.74) | 22.08 (7.90) |
| **FVC (L)** | 11 | 3.69 (1.04) | 3.76 (1.09) | 3.84 (1.12) | 3.58 (1.14) | 3.11 (1.15) | 3.75 (0.80) | 3.60 (1.14) |
| **FEV<sub>1</sub> (L)** | 11 | 2.92 (0.82) | 2.97 (0.91) | 2.90 (0.88) | 2.72 (0.99) | 2.11 (0.89) | 2.68 (0.80) | 2.70 (0.95) |
| **FEV/FVC (%)** | 11 | 79.77 (9.32) | 78.74 (8.48) | 75.69 (8.26) | 76.14 (12.69) | 68.61 (13.50) | 71.15 (12.58) | 75.18 (11.19) |
| **Mediastinal LA** | 9 | 12 (27.9%) | 37 (55.2%) | 61 (64.2%) | 12 (27.3%) | 47 (61.8%) | 1 (33.3%) | 170 (51.8%) |
| **Hilar LA** | 9 | 8 (18.6%) | 27 (40.3%) | 55 (57.9%) | 8 (18.2%) | 42 (55.3%) | 1 (33.3%) | 141 (43.0%) |
| **Micronodules** | 9 | 14 (32.6%) | 38 (56.7%) | 81 (85.3%) | 32 (72.7%) | 65 (85.5%) | 0 (0.0%) | 230 (70.1%) |
| **Conglomerate** | 9 | 0 (0.0%) | 2 (3.0%) | 26 (27.4%) | 8 (18.2%) | 53 (69.7%) | 0 (0.0%) | 89 (27.1%) |
| **Architectural Distortion** | 9 | 2 (4.7%) | 8 (11.9%) | 47 (49.5%) | 16 (36.4%) | 73 (96.1%) | 0 (0.0%) | 146 (44.5%) |
| **Traction Bronchiectasis** | 9 | 2 (4.7%) | 4 (6.0%) | 34 (35.8%) | 11 (25.0%) | 71 (93.4%) | 0 (0.0%) | 122 (37.2%) |
| **Fibrotic Abnormality** | 9 | 2 (4.7%) | 5 (7.5%) | 44 (46.3%) | 14 (31.8%) | 74 (97.4%) | 0 (0.0%) | 139 (42.4%) |
| **Ground-glass** | 9 | 5 (11.6%) | 16 (23.9%) | 36 (37.9%) | 15 (34.1%) | 52 (68.4%) | 0 (0.0%) | 124 (37.8%) |
| **Reticular Abnormality** | 9 | 1 (2.3%) | 11 (16.4%) | 44 (46.3%) | 19 (43.2%) | 60 (78.9%) | 0 (0.0%) | 135 (41.2%) |
| **Mosaic Attenuation** | 9 | 1 (2.3%) | 8 (11.9%) | 14 (14.7%) | 9 (20.5%) | 32 (42.1%) | 0 (0.0%) | 64 (19.5%) |

\*Includes subjects who both did not identify as Hispanic and also either identified as Asian, American Indian, Alaska Native, or did not identify as a single primary race.

### 3.2. Feature robustness and association with scanner

Among the 12 unharmonized datasets, use of unregistered images or empirical variogram data was typically associated with stronger associations between features and scanner model; see Figure 2. Matérn data were typically least associated with scanner and exhibited near uniform distributions of p-values for association with scanner model consistent with lack of association. R squared values were typically below 0.15. Harmonized datasets exhibited feature R squared values of approximately 0 and p-values biased towards 1 as expected.

**Figure 2.**
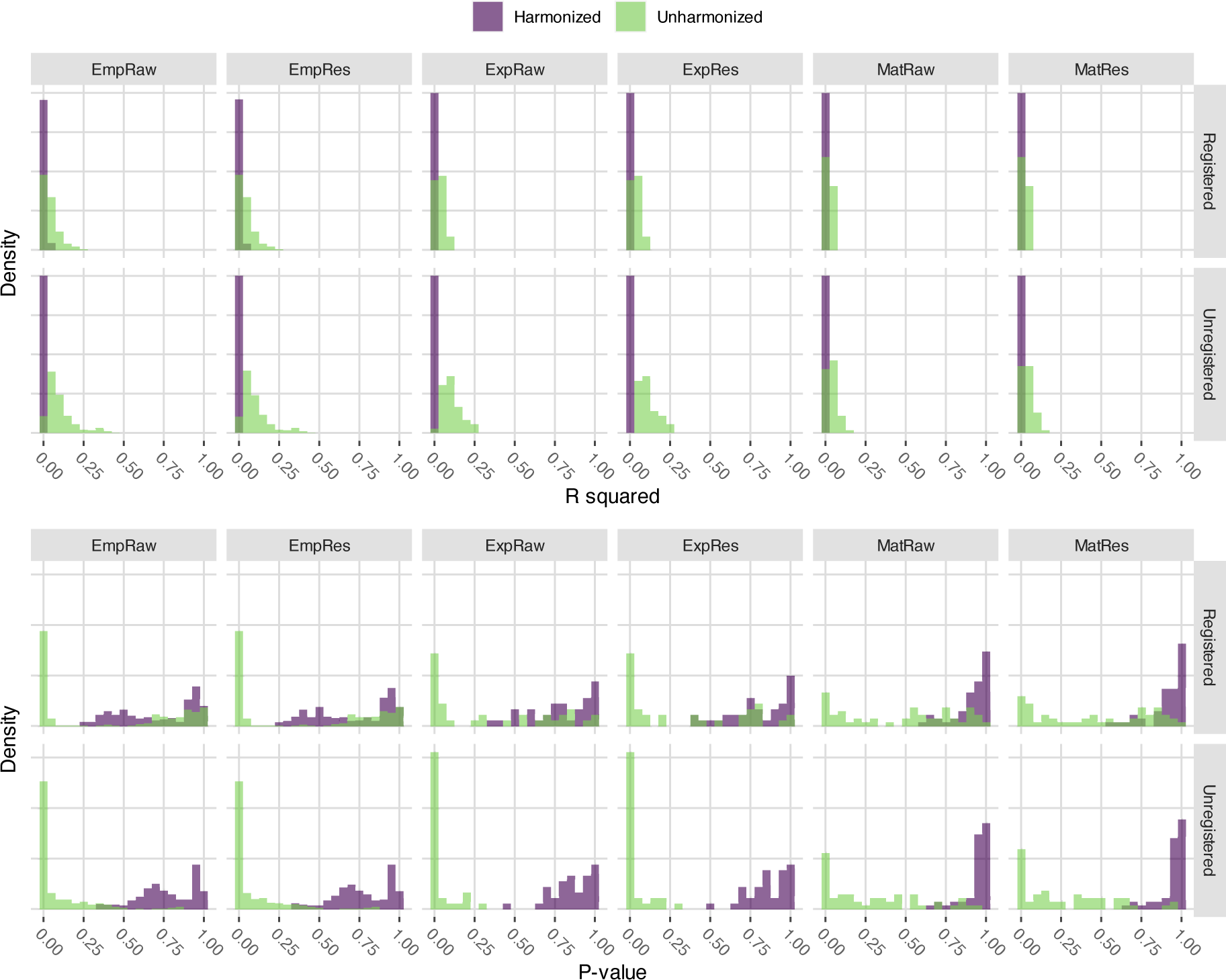
Density histograms of R squared values and p values obtained by regressing each engineered feature onto scanner model, stratified by the 24 engineered datasets (see Figure 1).

Pairs of matched variables from datasets differing only in handling of drift and use of harmonization typically exhibited excellent robustness (ICC*>* 0.9); see Figure 3, top two rows. Pairs of matched variables differing in use of registration (bottom row) exhibited moderate to good robustness for empirical datasets (0.5 *<*ICC*<* 0.9) and variable robustness for exponential and for Matérn datasets. Pairs of matched variables from datasets differing in harmonization exhibited slightly reduced ICC but harmonization was not substantially impactful; see Supplementary Figure 8. Spearman’s correlation was generally consistent with ICC, suggesting non-excellent robustness related to registration was generally not attributable to monotonic transformation; see Supplementary Figure 9.

**Figure 3.**
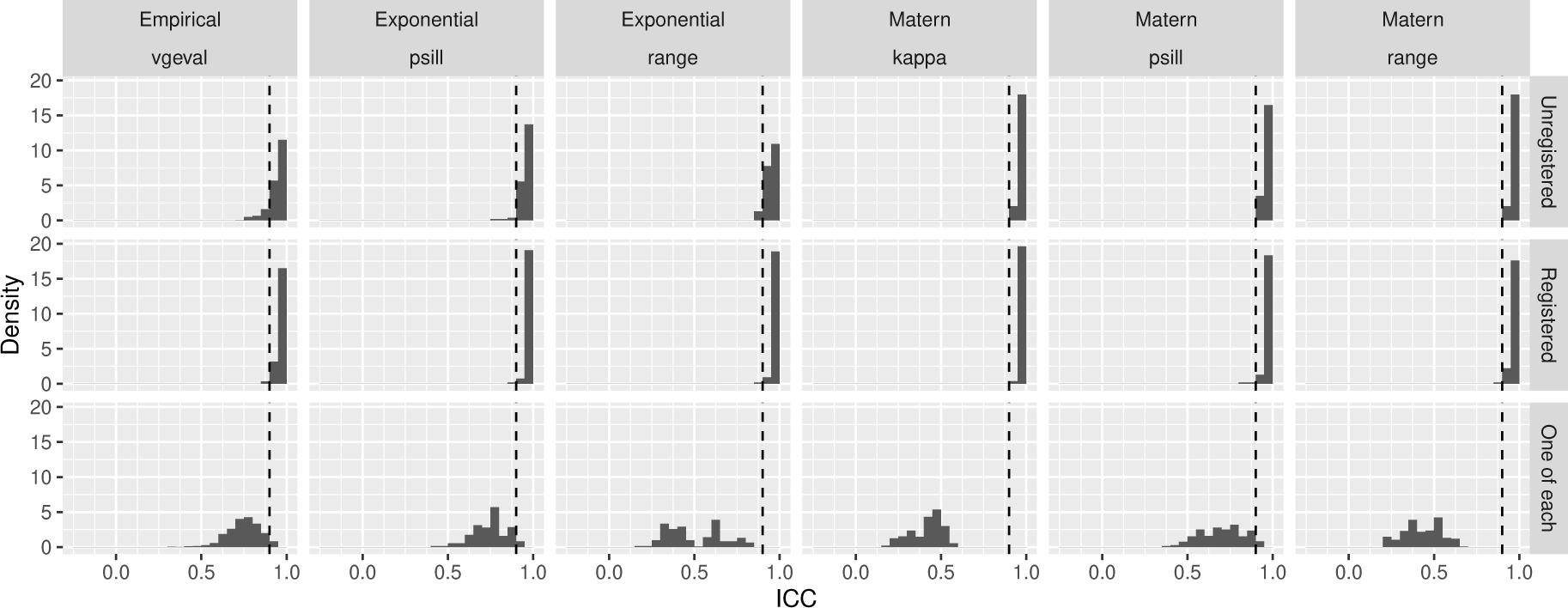
Histograms of intraclass correlation coefficients between matched variable pairs from different engineered datasets, stratified by engineered dataset (see Figure 1). Harmonized and unharmonized data are similar and thus pooled.

Matched variable pairs for which one variable was from Matérn data and the other was from exponential data exhibited differing robustness patterns by parameter; see Supplementary Figure 10. Patterns for the psill parameter were consistent with patterns described above, with robustness typically excellent unless datasets differed by registration as well. Robustness for the range parameter was highly variable and especially poor when datasets differed by registration, suggesting these data may not be comparable.

### 3.3. Cluster results, consistency, and association with scanner

The 48 clustering analyses typically resulted in between 6 and 8 clusters; see Supplemental Table 3. Most cluster analyses selected models which were not sparse; see Supplemental Figure 11. Analyses of empirical variogram data selected all variables. Analyses of fit Matérn variogram model parameters, especially those using MICL for model selection, tended to be sparser. Partial sill parameters were deemed relevant from all locations by all analyses while range exhibited variable relevance and kappa parameters were typically deemed relevant almost everywhere for analyses using BIC, and relevant at the bottom of the lungs for analyses using MICL. Relevance patterns did not differ substantially between left and right lungs. Analyses of fit exponential variogram model parameters similarly deemed partial sill parameters to be relevant everywhere. Range parameters were deemed relevant everywhere when registration was used in feature engineering and at the tops and bottoms of the lung when it was not. Harmonization and drift were not substantial factors for relevance.

All cluster analyses from harmonized data were not significantly associated with scanner (*p >* 0.1), as expected; see Table 2. Cluster analyses from unharmonized, unregistered data exhibited stronger, more significant associations (*p <* 0.001) with scanner model than cluster analyses from unharmonized, registered data (*p >* 0.01), consistent with patterns of associations between variogram features and scanner model. Association strength between clusters from unharmonized, unregistered data and scanner model ranged from 0.15 to 0.3 as measured by Cramér’s V, indicating cluster analyses did differ substantially from scanner model.

**Table 2.** For each of the 48 cluster analyses (see Figure 1), we report the strength of association between clusters and scanner model using Cramér’s V. In parentheses, we report simulated Fisher’s exact test p-values (20,000 replicates) with * indicating *p <* 0.001 and - indicating *p >* 0.1.

| Data | Drift | IC | Registered |  | Unregistered |  |
| --- | --- | --- | --- | --- | --- | --- |
|  |  |  | Harmonized | Unharmonized | Harmonized | Unharmonized |
| Empirical | Linear | BIC | 0.000 (-) | 0.086 (0.035) | 0.055 (-) | 0.188 (*) |
| Empirical | Linear | MICL | 0.000 (-) | 0.092 (0.034) | 0.000 (-) | 0.163 (*) |
| Empirical | Raw | BIC | 0.062 (-) | 0.078 (0.050) | 0.020 (-) | 0.178 (*) |
| Empirical | Raw | MICL | 0.000 (-) | 0.126 (0.005) | 0.000 (-) | 0.151 (*) |
| Exponential | Linear | BIC | 0.023 (-) | 0.069 (0.059) | 0.000 (-) | 0.271 (*) |
| Exponential | Linear | MICL | 0.065 (-) | 0.052 (-) | 0.000 (-) | 0.279 (*) |
| Exponential | Raw | BIC | 0.057 (-) | 0.083 (0.013) | 0.000 (-) | 0.247 (*) |
| Exponential | Raw | MICL | 0.000 (-) | 0.076 (0.035) | 0.000 (-) | 0.276 (*) |
| Matern | Linear | BIC | 0.000 (-) | 0.000 (-) | 0.000 (-) | 0.189 (*) |
| Matern | Linear | MICL | 0.000 (-) | 0.092 (0.012) | 0.000 (-) | 0.187 (*) |
| Matern | Raw | BIC | 0.000 (-) | 0.076 (0.076) | 0.000 (-) | 0.188 (*) |
| Matern | Raw | MICL | 0.000 (-) | 0.031 (-) | 0.000 (-) | 0.196 (*) |

Pairwise comparisons of cluster analyses using Cramér’s V showed strong consistency across analyses; see Figure 4. All comparisons produced values above 0.4, 40% produced values above 0.6, and 15% produced values above 0.7. 70% of comparisons of unregistered analyses produced values above 0.6 while 60% of comparisons of registered analyses produced values above 0.6. 48% of comparisons of unharmonized analyses produced values above 0.6 while 54% of comparisons of harmonized analyses produced values above 0.6.

**Figure 4.**
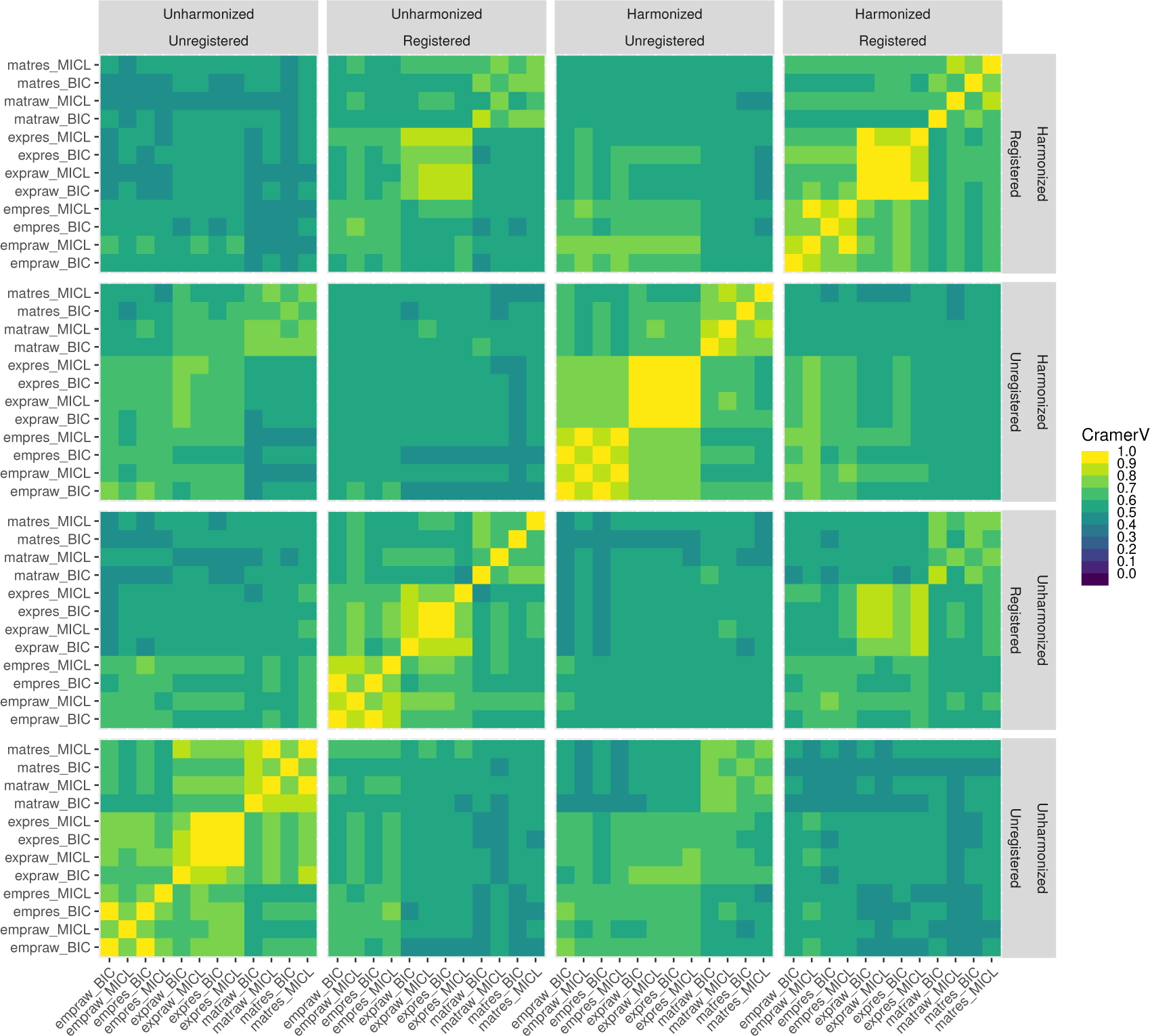
Pairwise comparisons by Cramér’s V of 48 cluster analyses (see Figure 1).

Registration was the most influential factor in consistency across the 48 analyses, followed by harmonization and data type (empirical, exponential, or Matérn). Strongest consistency was observed between analyses differing only in handling of drift or selection criterion.

### 3.4. Clinical outcomes and demographics by clusters

Comparisons of demographics, PFT, and VAS to each of the 48 clustering analyses are summarized; see Figure 5 for unadjusted association strength and Supplemental Figure 12 for corresponding p-values.

**Figure 5.**
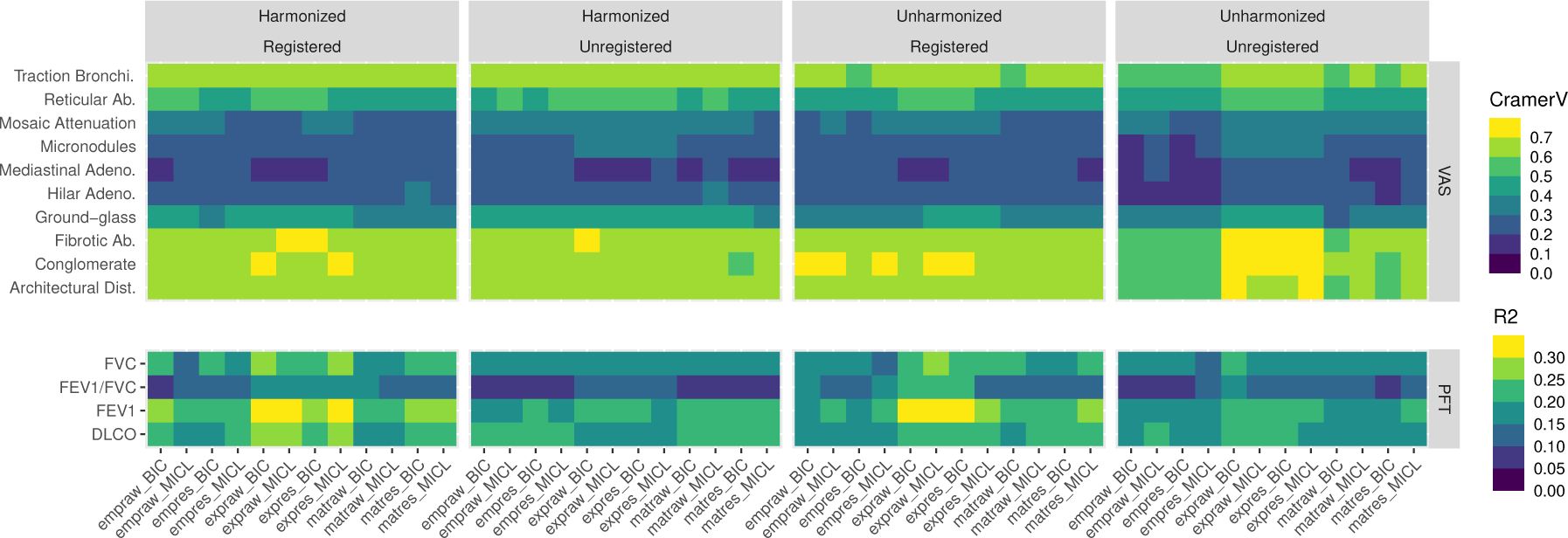
Cramér’s V comparing 48 cluster analyses (see Figure 1) to VAS and linear model r-squared values predicting PFT from each analysis without adjustment.

All analyses produced clusters which differed significantly by DLCO, FEV1, and FVC (*p <* 0.001) and by BMI and FEV1/FVC (*p <* 0.05). Most analyses produced clusters which differed significantly by self-reported race and ethnicity indicator (*p <* 0.001), with analyses from data not harmonized for scanner effects exhibiting stronger association. Most analyses produced clusters which differed significantly by sex (*p <* 0.05), though no clusters produced from unharmonized empirical variogram data differed significantly by sex. Most analyses produced clusters which did not differ significantly by age and height (*p >* 0.05), though all analyses from registered exponential data differed significantly by height (*p <* 0.001).

Most analyses produced clusters which differed significantly (*p <* 0.001) by each VAS measure except mediastinal lymphadenopathy.

A comparison of cluster analyses with VAS using Cramér’s V showed that clusters tended to capture architectural distortion, conglomerate mass, fibrotic abnormality, and traction bronchiectasis particularly well (typical Cramér’s V *>* 0.6), as well as ground-glass, mosaic attenuation, and reticular abnormality more modestly (typical Cramér’s V *>* 0.3); see Figure 5. A comparison of cluster analyses with PFT further suggested strong associations between PFT and clusters. Clusters generally explained 15-35% of variability observed in FEV1, FVC, and DLCO. Clusters obtained from fitting exponential variogram models to registered data were particularly strongly associated with FEV1 and typically explained 25-35% of variability. Overall, clusters obtained from registered data exhibited slightly stronger unadjusted associations with PFT outcomes than those from unregistered data.

### 3.5. Cluster associations with clinical outcomes after adjustment

After adjusting for base demographic factors, clusters exhibited strong associations with PFT; see Figure 6 for R squared summaries, Supplemental Figure 13 for p-value summaries. Clusters typically explained another 10-15% of variability beyond base adjustment factors.

**Figure 6.**
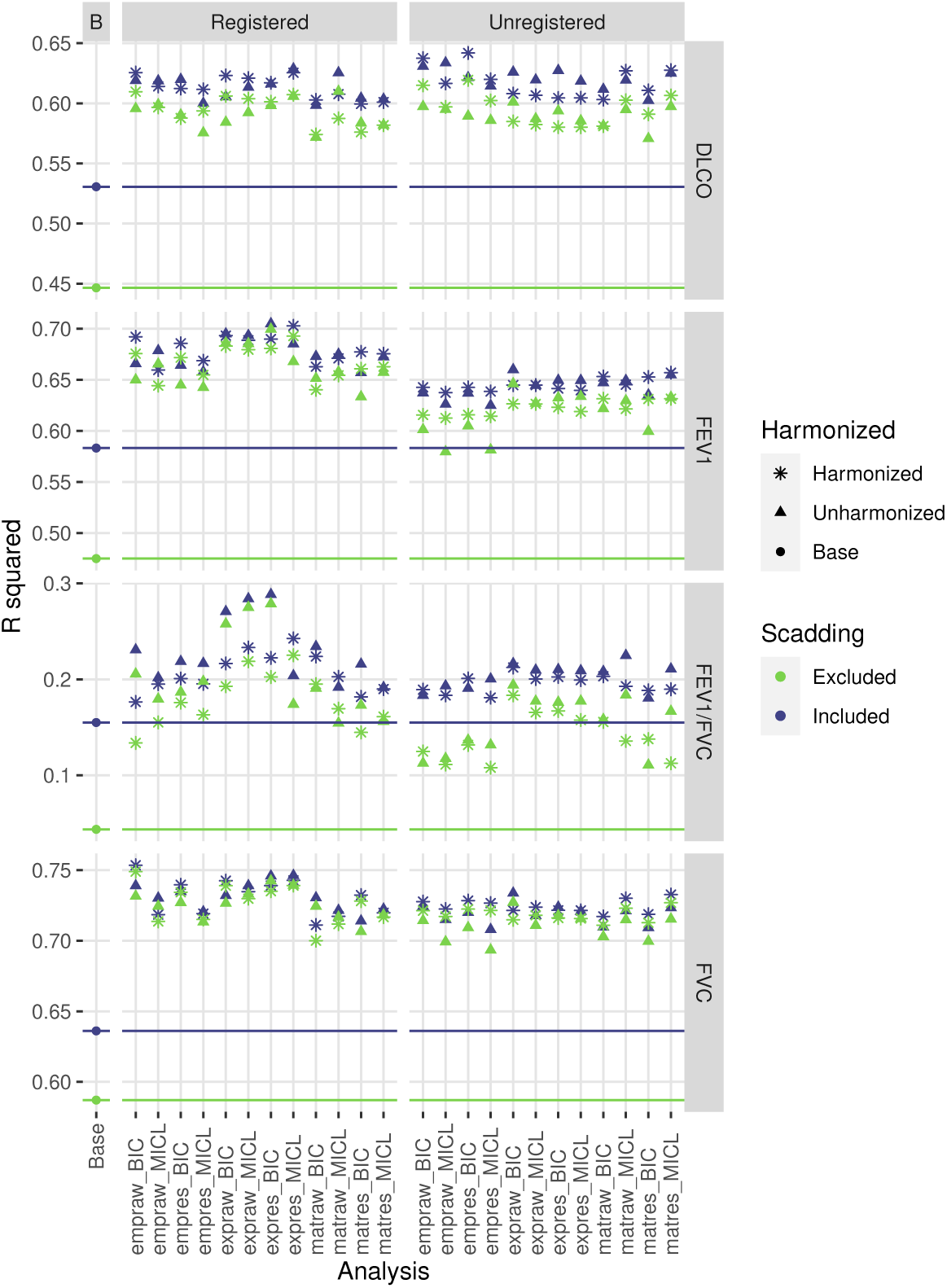
R squared values for various linear models predicting PFT from base adjustment factors, cluster analysis grouping, and Scadding stage, with color indicating inclusion or exclusion of Scadding stage as a variable and shape indicatating whether engineered data were harmonized prior to clustering. For each of 48 cluster analyses (see Figure 1), we fitted a model with base factors and group and a model with base factors, group, and Scadding. Additionally, we fitted a model with base factors and a model with base factors and Scadding, indicated as the base analyses. For reference, horizontal lines indicate R squared values for base models.

Associations with DLCO, FEV1, and FVC were similar compared to each other and across cluster analyses. Scadding stage typically lost significance (*p >* 0.001 or *p ≈* 0.001) in the presence of clusters and clusters were significant (*p* ≪ 0.001) with or without adjusting for Scadding. Clusters alone explained much more variability than Scadding stage alone beyond base adjustment factors. Scadding stage did not explain substantially more variation beyond cluster grouping and base adjustment factors. Clusters from registered data typically exhibited slightly stronger associations, with the effect of registration most pronounced for FEV1.

For FEV1/FVC, clusters produced from registered data exhibited stronger associations more often with or without adjusting for Scadding stage, especially when clusters were produced from unharmonized exponential data. Clusters alone typically explained as much variability as Scadding stage alone beyond base adjustment factors.

Most full models of VAS produced (training) AUC roughly 0.05 greater than Scadding models, while most cluster models produced AUC roughly the same as Scadding models; see Figure 7 for AUC summaries, Supplemental Figure 14 for p-value summaries. Full models of architectural distortion, conglomerate mass, fibrotic abnormality, and traction bronchiectasis all produced AUC around or above 0.9, with full models of ground-glass, micronodules, mosaic attenuation, and reticular abnormality producing AUC around or above 0.8. All models of hilar and mediastinal lymphadenopathy failed to produce AUC above 0.75.

**Figure 7.**
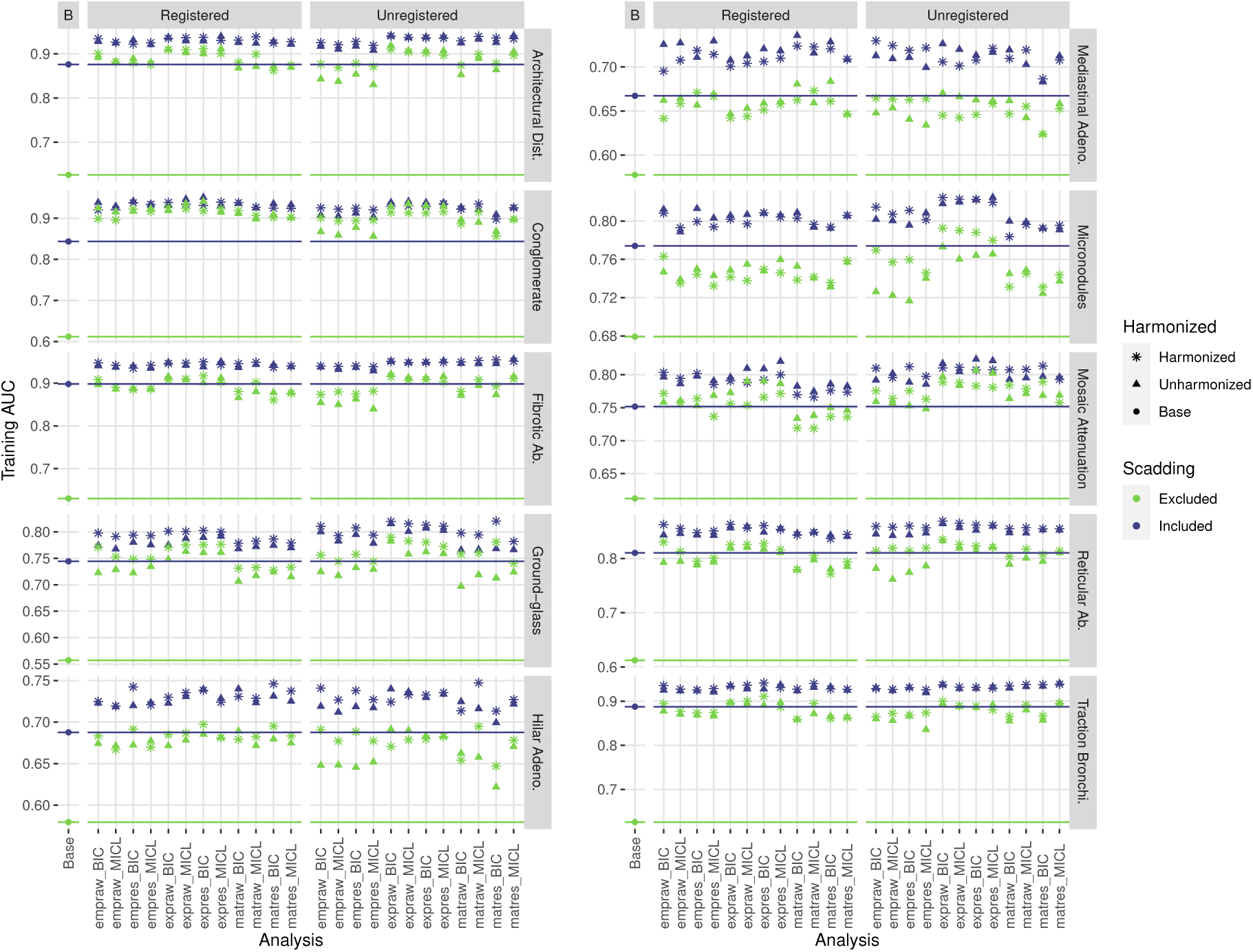
AUC values for various Firth-type logistic models predicting VAS from base adjustment factors, cluster analysis grouping, and Scadding stage, with color indicating inclusion or exclusion of Scadding stage as a variable and shape indicatating whether engineered data were harmonized prior to clustering. For each of 48 cluster analyses (see Figure 1), we fitted a model with base factors and group and a model with base factors, group, and Scadding. Additionally, we fitted a model with base factors and a model with base factors and Scadding, indicated as the base analyses. For reference, horizontal lines indicate AUC values for base models.

AUC values were typically comparable between registered and unregistered analyses, and between harmonized and unharmonized analyses.

Association between clusters and VAS measures was typically significant (*p <* 0.001) for cluster and full models of architectural distortion, conglomerate mass, fibrotic abnormality, reticular abnormality, and traction bronchiectasis. Associations between clusters and VAS measures were typically significant in cluster models but lost significance after adjustment for Scadding stage in models of ground-glass, hilar adenopathy, micronodules, and mosaic attenuation. Scadding stage typically remained significant in full models of architectural distortion, fibrotic abnormality, hilar and mediastinal lymphadenopathy, micronodules, reticular abnormality, and traction bronchiectasis, while dropping to marginal significance (*p ≈* 0.001) in full models of conglomerate mass and ground-glass and losing significance (*p >* 0.001) in full models of mosaic attenuation.

## 4. Discussion

In this paper, we investigated potential clinical relevance of variograms-based feature engineering pipelines for quantification of chest CT imaging and phenotyping in the study of sarcoidosis. Factors of interest included feature robustness, consistency of identified phenotypes across approaches, and strength of association between identified phenotypes and clinical measures of disease.

Consistent with the literature, computed variogram-based features were not robust to the decision to register images to a template (frequently ICC *<* 0.5). If the decision on registration was fixed, feature robustness was typically excellent with respect to other decisions (ICC*>* 0.9). Even in absence of robust feature computation, however, resulting clusters from each of 48 clustering analyses were surprisingly consistent (typical Cramér’s V *>* 0.5). Mild but statistically significant associations between features and scanner model used in image acquisition were found for all 24 unharmonized datasets, especially empirical and exponential datasets, which persisted through to associations between phenotypes and scanner model. Associations between features and scanner were mild at worst, unlike in radiomics (Rizzo et al. 2018, Mackin et al. 2015). Registering images before feature extraction appeared to reduce feature and phenotype association with scanner model.

Presence and strength of disease signal were impressively consistent across all 48 approaches considered for processing images, engineering features, and phenotyping/clustering. Associations of variogram-based phenotypes with PFT were observed to be similarly strong compared to previously identified associations of radiomics-based phenotypes with PFT in the GRADS cohort (Carlson et al. 2024) and results were consistent with similar radiomics-based studies of PFT and VAS in lung diseases such as COPD and ILD (Occhipinti et al. 2019, Schniering et al. 2022). Associations of variogram-based phenotypes with visual assessments of architectural distortion, conglomerate masses, fibrotic abnormalities, and traction bronchiectasis (optimistic AUC ≈ 0.9) establish strong potential relevance of variogram-based approaches in future study of sarcoidosis and quantitative biomarker development. Lowest strength associations between clusters and measures of disease were observed for lymphadenopathy measures (optimistic AUC between 0.65 and 0.75), though we note that lymph nodes were masked out of images for all 48 pipelines.

Consistency in identified clusters and disease signal across approaches suggested that use of unharmonized variogram data from unregistered images, consistent with a simple, expedient, and generalizable processing pipeline, is reasonable for future study of sarcoidosis. Further, the low-dimension variogram-based feature sets considered here are more interpretable and apparently much less influenced by scanner model than radiomics while also showing strong association with disease features. This work suggests variogram-based features are a promising alternative to classic radiomics in sarcoidosis.

### 4.1. Limitations

The GRADS study enrolled subjects representing clinically pre-defined phenotypes, and so analyses of heterogeneity in a subcohort may not be generalizable to clinical populations. Furthermore, CT scans obtained for the GRADS study were more consistent in image acquisition and reconstruction than is typical for clinical populations. Variation in image acquisition protocol could not be assessed. Future investigations should more thoroughly assess pipeline robustness to variation in image acquisition and reconstruction, as well as the highly related factor of image resolution.

## 5. Conclusion

All feature engineering pipelines and clustering approaches considered produced similar measures of heterogeneity and strong association with sarcoidosis disease signal as measured by PFT and VAS, clearly establishing potential for clinical relevance of variogram-based approaches in future research. While clusters produced from unregistered data were typically more significantly associated with scanner model than those produced from registered data, this association was mild in strength and did not appear to impede detection of disease related signal in subsequent analyses. As such, optimality of pipelines and approaches among those considered might reasonably be selected on other bases, such as simplicity and expediency of pipeline, dimensionality of data produced, robustness of feature computation, and appropriateness of pipelines for clinical cohorts having more variability in CT image metadata such as original reconstructed resolution, convolution kernel used in reconstruction, and retention of axial slices.

In particular, future investigation of variogram-based features in clinical sarcoidosis cohorts might reasonably consider unharmonized, unregistered Matérn data without accounting for drift, corresponding with a balance of simplicity in implementation and robustness of engineered features. In principle, this harmonization- and registration-free pipeline should also be more readily applicable to both research and clinical cohorts without substantial modification. Building and applying a template is typically a time-consuming cohort-specific process with the potential to impede generalizability while harmonization directly impedes generalization to new contexts.

Registered image data do appear slightly more appropriate for establishing clinically relevant phenotypes through clustering as registration seems to reduce association of engineered features and clustering-based phenotypes with scanner model; however, care would need to be taken to ensure templates and registration process were appropriate for application to diverse clinical populations of sarcoidosis patients in support of generalizable research.

## Data Availability

GRADS data are available via the process laid out by the GRADS consortium.

## Acknowledgments

This work was supported by the National Institutes of Health under Grants R01 HL114587, R01 HL142049, R01 HL152735, and T32 HL007085.

Data from the GRADS study was supported under Grants U01 HL112707, U01 HL112707, U01 HL112694, U01 HL112695, U01 HL112696, U01 HL112702, U01 HL112708, U01 HL112711, U01 HL112712.

## Supplemental Figures and Tables

**Figure 8.**
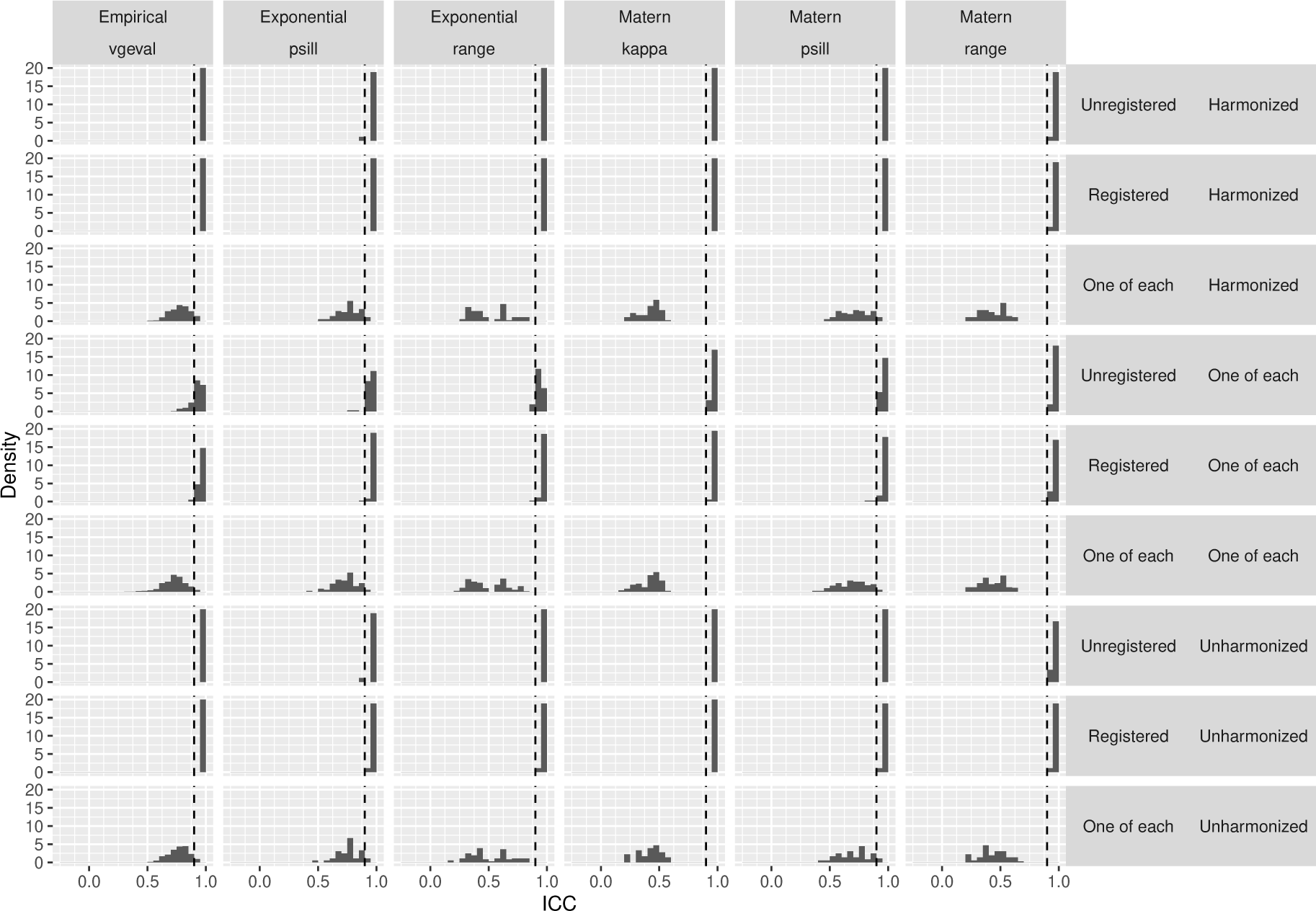
Histograms of intraclass correlation coefficients between matched variable pairs from 24 different engineered datasets (see Figure 1).

**Figure 9.**
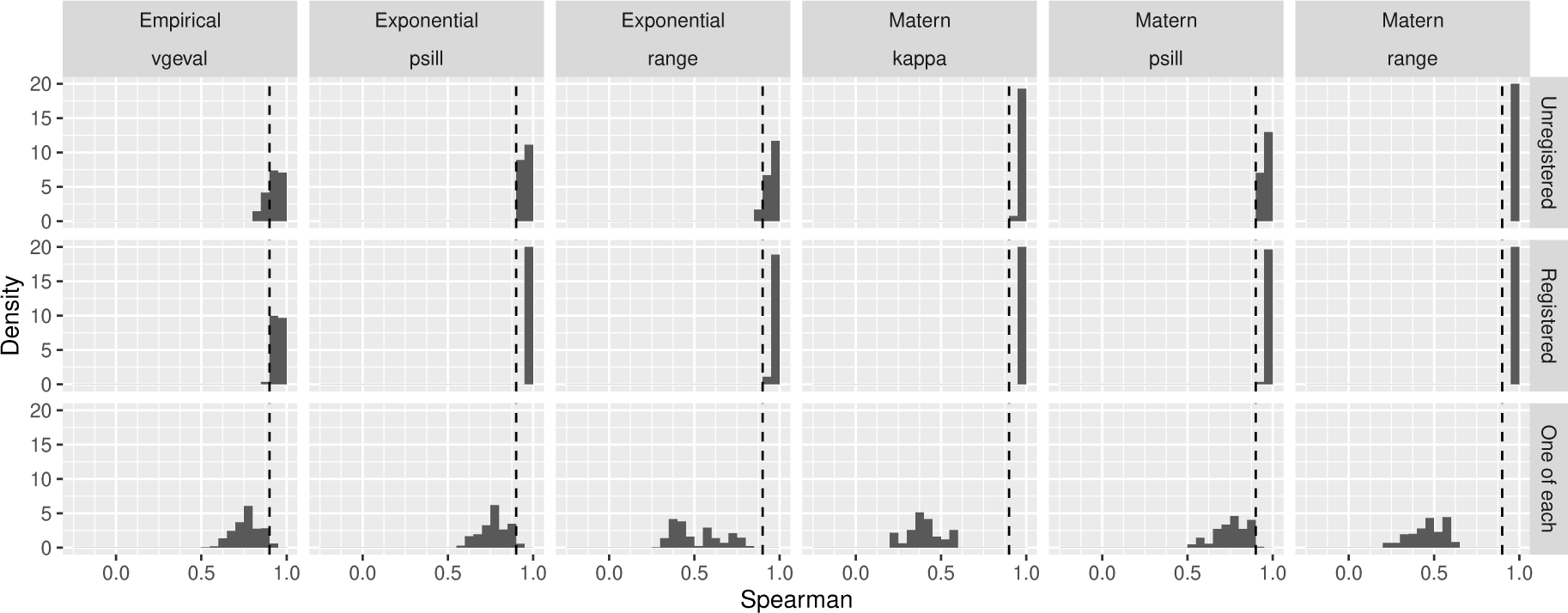
Histograms of Spearman’s correlation between matched variable pairs from 24 different engineered datasets (see Figure 1). Harmonized and unharmonized data are similar and thus pooled.

**Figure 10.**
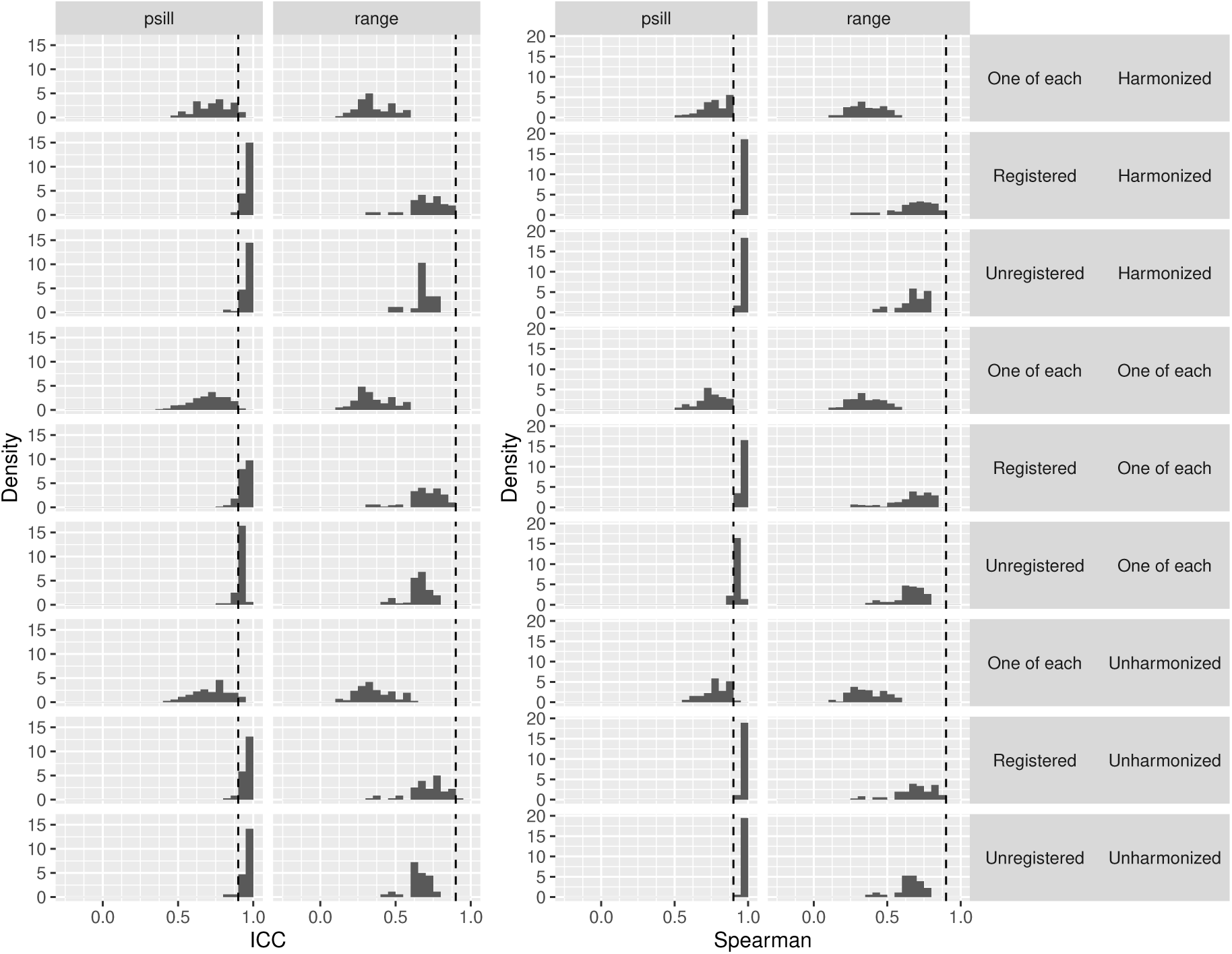
Histograms of intraclass correlation coefficients and Spearman’s correlation between matched variable pairs, one each from exponential and Matérn data, stratified by type of feature engineered (partial sill or range), registration (both datasets from registered data, both from unregistered data, or one of each), and harmonization (both datasets from harmonized data, both from unharmonized data, or one of each).

**Table 3.**
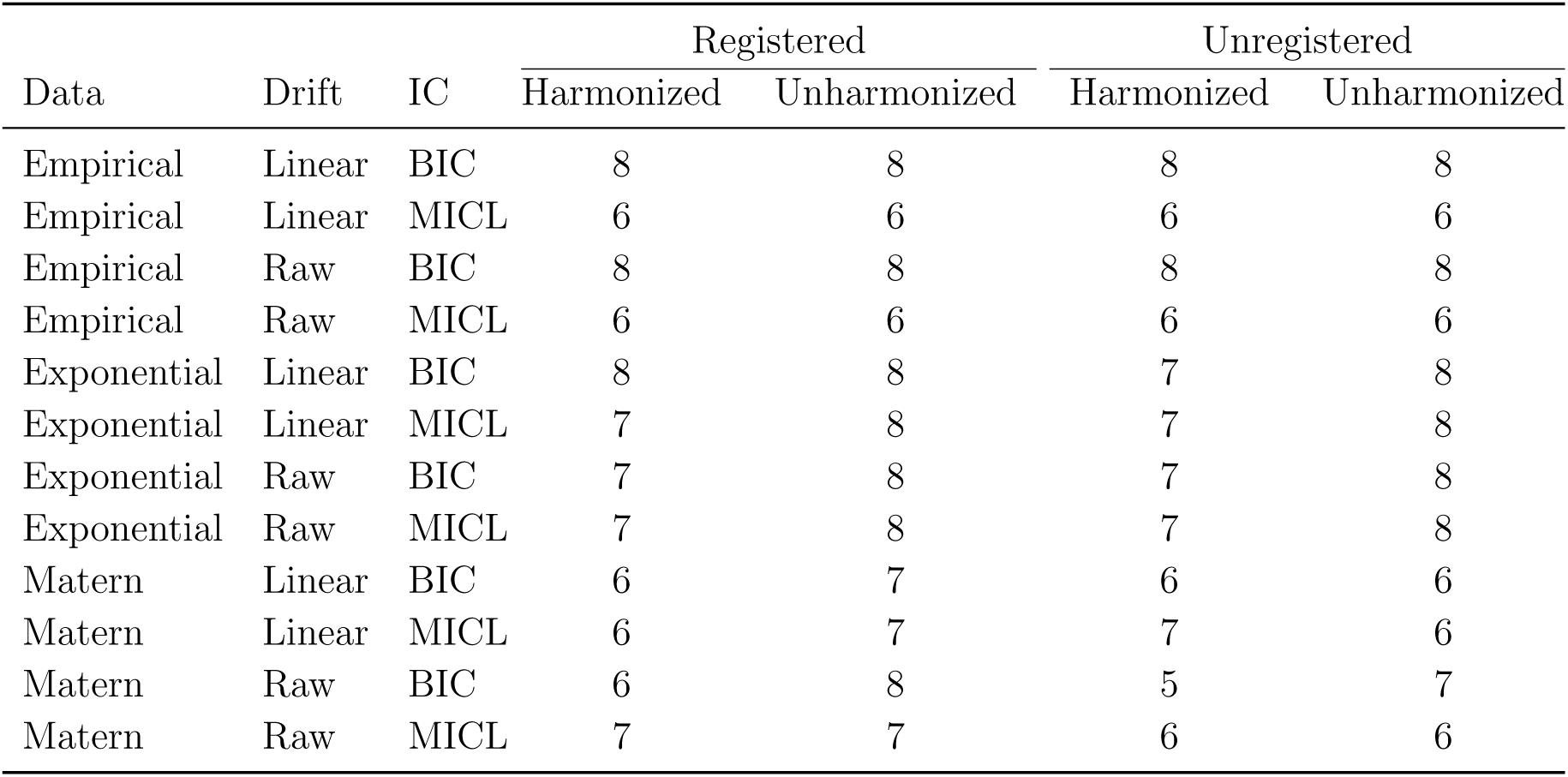
For each of 48 cluster analyses (see Figure 1), we report the number of clusters selected.

| Data | Drift | IC | Registered |  | Unregistered |  |
| --- | --- | --- | --- | --- | --- | --- |
|  |  |  | Harmonized | Unharmonized | Harmonized | Unharmonized |
| Empirical | Linear | BIC | 8 | 8 | 8 | 8 |
| Empirical | Linear | MICL | 6 | 6 | 6 | 6 |
| Empirical | Raw | BIC | 8 | 8 | 8 | 8 |
| Empirical | Raw | MICL | 6 | 6 | 6 | 6 |
| Exponential | Linear | BIC | 8 | 8 | 7 | 8 |
| Exponential | Linear | MICL | 7 | 8 | 7 | 8 |
| Exponential | Raw | BIC | 7 | 8 | 7 | 8 |
| Exponential | Raw | MICL | 7 | 8 | 7 | 8 |
| Matern | Linear | BIC | 6 | 7 | 6 | 6 |
| Matern | Linear | MICL | 6 | 7 | 7 | 6 |
| Matern | Raw | BIC | 6 | 8 | 5 | 7 |
| Matern | Raw | MICL | 7 | 7 | 6 | 6 |

**Figure 11.**
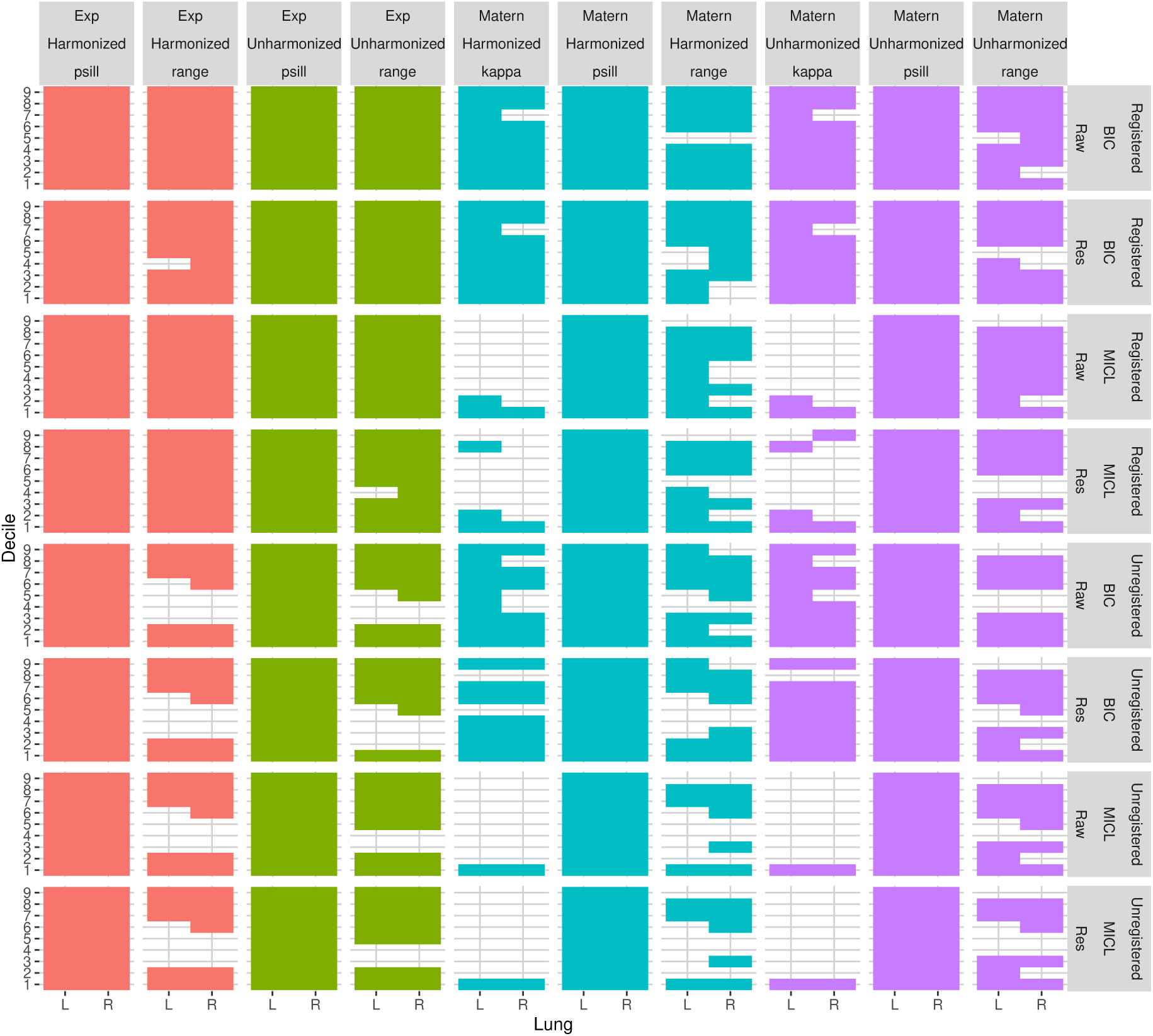
Relevant variables selected in producing clusters in each of 32 cluster analyses of model fit variogram data (see Figure 1). An additional 16 cluster analyses of empirical variogram data each deemed all variables relevant and are not depicted. Variables are indicated as a combination of lung decile (1-9), lung (left or right), and feature type (psill, range, or kappa).

**Figure 12.**
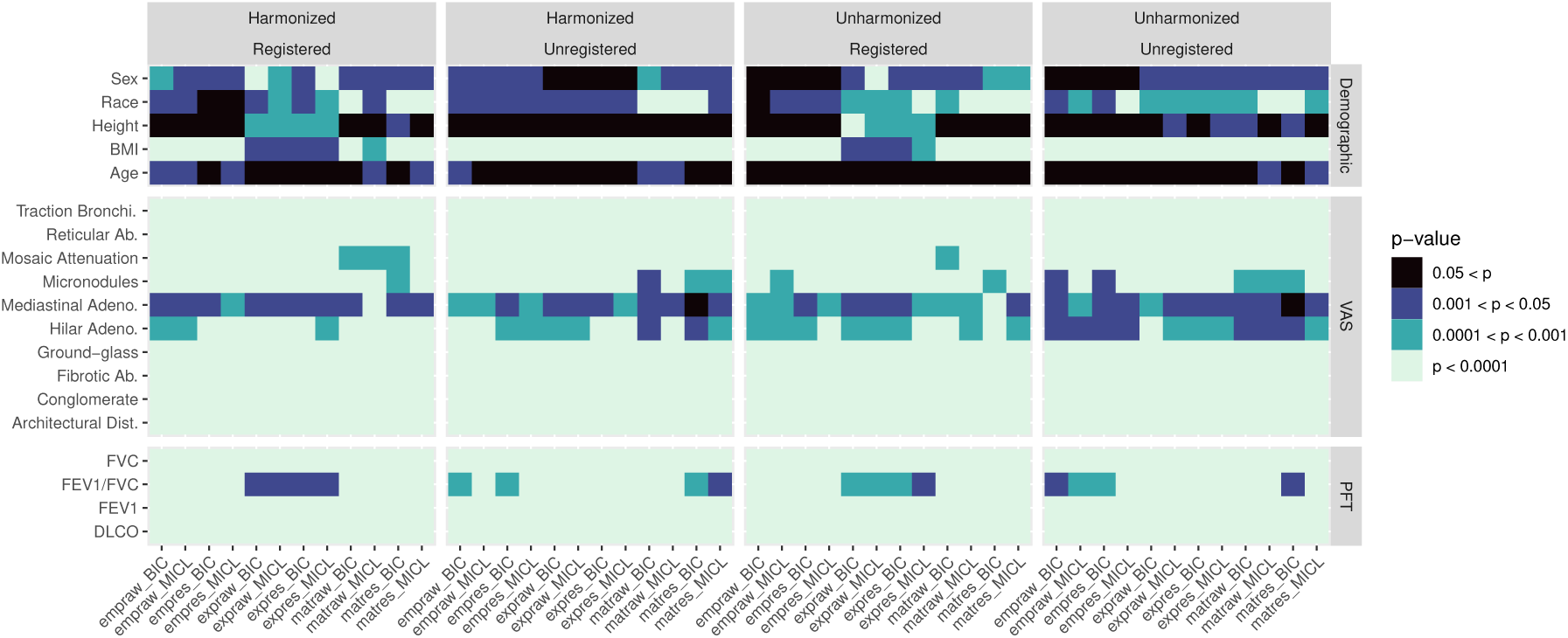
P-values for significance of association in comparing each of 48 cluster analyses (see Figure 1) to demographics, VAS, and PFT without adjustment. Fisher’s exact test was used for VAS measures, sex, and self-identified race while Welch’s ANOVA was used for PFT measures, age, height, and BMI.

**Figure 13.**
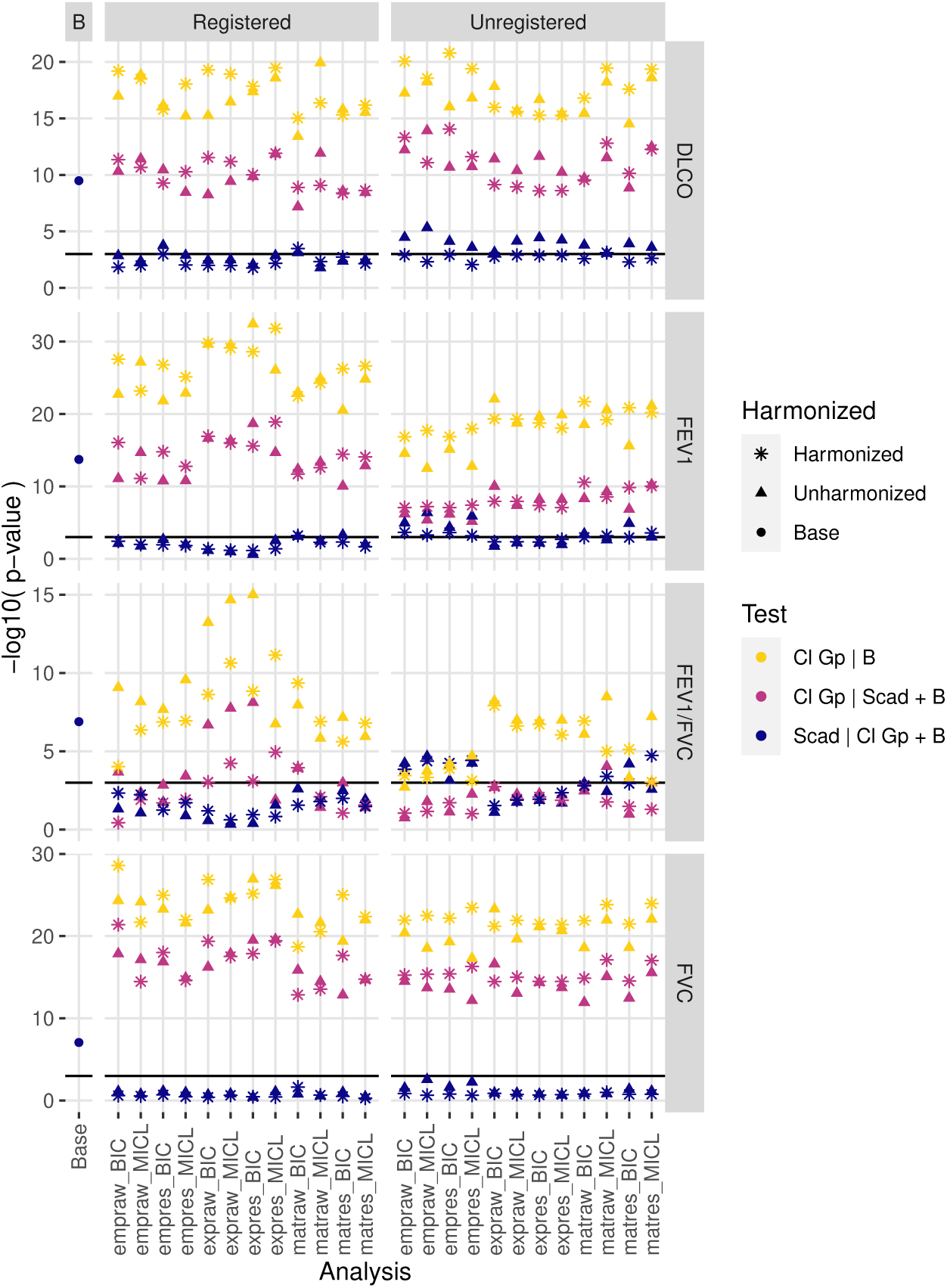
Likelihood ratio test p-values comparing various linear models predicting PFT from base adjustment factors, cluster analysis grouping, and Scadding stage after negative log10 transformation. For each cluster analysis, a model with base factors and group was compared to the base model, a model with base factors, group, and Scadding was compared to the model with base factors and Scadding, and a model with base factors, group, and Scadding was compared to the model with base factors and group. Additionally, the model with base factors and Scadding was compared to the model with base factors, indicated as the base analysis. A black horizontal line indicates a p-value of 0.001, with higher plotted values corresponding with lower, more significant p-values.

**Figure 14.**
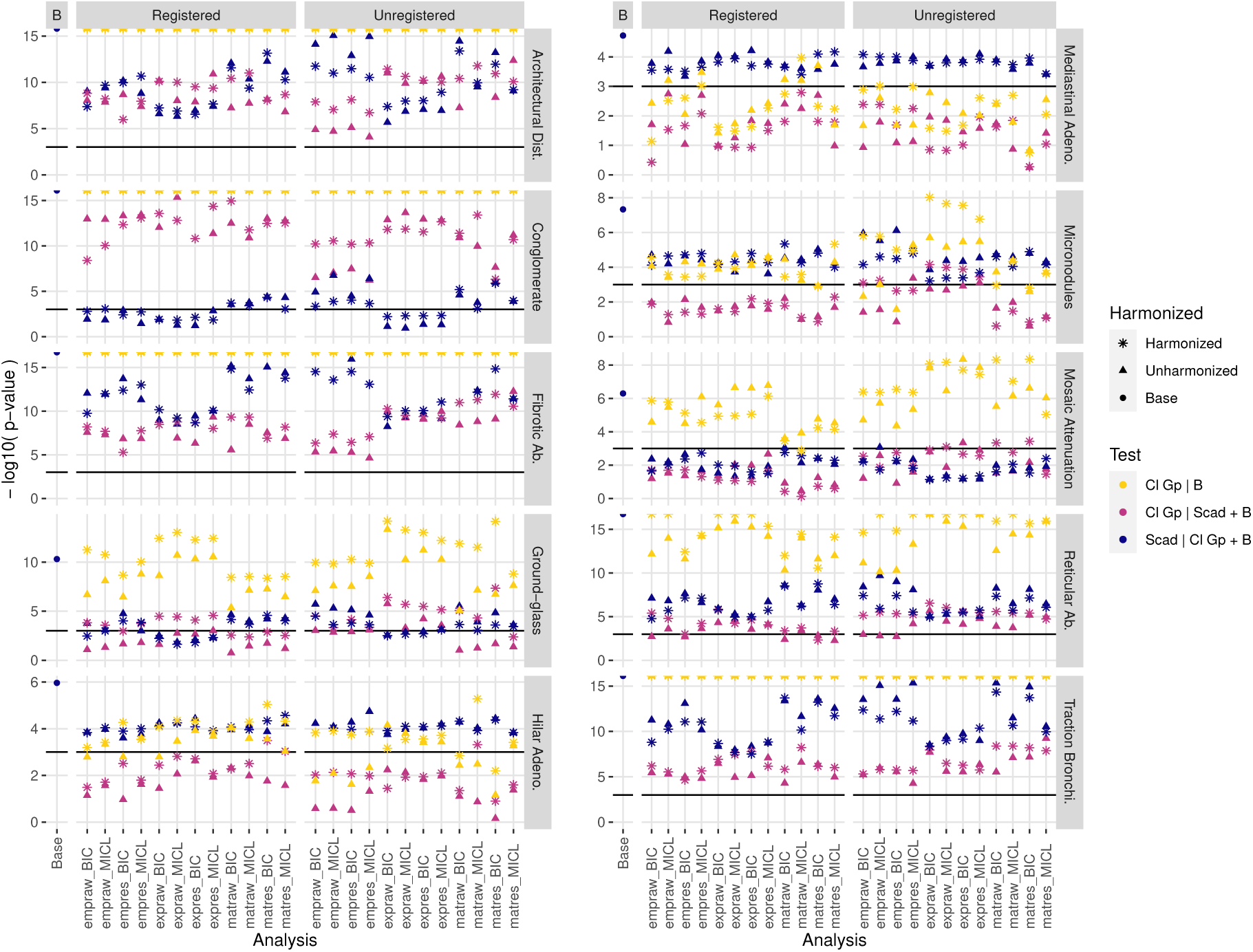
Penalized likelihood ratio test p-values comparing various Firth-type logistic models predicting VAS from base adjustment factors, cluster analysis grouping, and Scadding stage after negative log10 transformation. For each cluster analysis, a model with base factors and group was compared to the base model, a model with base factors, group, and Scadding was compared to the model with base factors and Scadding, and a model with base factors, group, and Scadding was compared to the model with base factors and group. Additionally, the model with base factors and Scadding was compared to the model with base factors, indicated as the base analysis. A black horizontal line indicates a p-value of 0.001, with higher plotted values corresponding with lower, more significant p-values. Apparent capping of p-values for significance of cluster group in some models is a result of near perfect or perfect separation producing p-values indistinguishable from 0 up to computer precision.

